# Efficient risk mitigation planning for a clinical trial

**DOI:** 10.1101/2022.11.01.22281727

**Authors:** Vladimir Shnaydman

## Abstract

Each clinical trial faces numerous risks. Neglecting or reacting spontaneously at each materialized risk may delay trial completion, affect trial objectives and even derail the clinical trial. At the same time, pre-allocating excessive contingency resources to eliminate all potential risks, even with a low probability of occurrence and impact, may not be necessary. Therefore, optimal risk planning means finding the “right” balance between contingency resources, risk mitigation strategy, and risk tolerance.

Most risk planning publications focus only on risk identification and risk assessment. Risk mitigation strategy planning is often overlooked. A contingency budget is routinely assigned as a percentage of a clinical trial budget, e.g., 10-20%, and is not aligned with the clinical trial risk level. Therefore, to derive an effective clinical trial risk mitigation strategy, a new methodology based on mathematical modeling was developed and validated for a Ph2 clinical trial.

## Introduction

Clinical trials are complex projects with numerous risks that could influence their outcome. Risks could be associated with potentially low enrolling sites, protocol amendments, adverse events, high dropout rate, clinical supply stock-out, biological sample handling, etc. Poor risk planning can delay or even compromise clinical trial results. Therefore, risk planning should be integral to risk-based quality management (RBQM) and clinical trial budgeting. If a risk event occurs during clinical trial execution without proper pre-allocated contingency resources, trial execution could be significantly delayed or even compromised.

The typical risk planning process includes several interconnected steps: identification of potential risks, risk assessment, risk response plan development, and risk monitoring.

### Risks identification

Potential risks could be identified via interviewing, risk registers of previous projects, etc.

### Risks assessment

Qualitative risk analysis will be used to evaluate potential risks in clinical trials due to the broad diversity of risks and the absence of relevant data for quantitative risk analysis^1^. Qualitative risk analysis includes subjective judgment of the risk as the probability of an unfortunate event and impact, typically on some numerical scale, e.g., from 1 to 5. The risk score is a product of the risk probability (or risk) and its impact. The technique is described in many sources, for example, in [1,2].

In [3, 4], it is recommended (nonbinding recommendation) to calculate risk assessment (Risk Priority Number - RPN) as a product of likelihood, impact, and detectability. In this paper, instead of risk detectability, mitigation costs are used. Deriving accurate mitigation costs assume a high level of detectability of each risk.

### The risk response plan

aims to reduce risk. Several options could be considered: risk avoidance (an alternative approach that does not include a particular risk), risk prevention, risk transfer (e.g., insurance to cover patients’ safety), risk acceptance, and risk mitigation.

In this paper, risk mitigation assumes that several mutually exclusive mitigation actions could be associated with each risk. For example, backup clinical sites could be identified in countries A or B. It allows a higher flexibility in risk mitigation strategy.

Although there is a consensus that developing risk mitigation plans is an integral part of clinical trial risk management, only a few mainly descriptive papers were published in the last few years [5-9]. The author could not find commonly accepted techniques, models, or analytical tools to support the derivation of effective risk mitigation strategies for a clinical trial. Most efforts in risk analytics focus on risk identification, assessment, and monitoring. A contingency budget is routinely assigned as a percentage of a clinical trial budget, e.g., 10-20%, and is not aligned with the clinical trial risk level.

At the same time, the risk mitigation planning problem was addressed in many papers applied to other industries, e.g., construction. The authors [10-19] developed sophisticated analytical tools from optimization to Monte Carlo simulation.

Optimization tools [10-14, 16] are used for budget/contingency budget allocation. Monte Carlo simulation tools are applied for risk mitigation analysis for project and portfolio planning, where critical risk metrics like project delay, cost of the delay, and lost revenue can be used. Calculating the risk impact regarding costs, time delay, and lost revenue is rewarding. However, deriving these parameters in clinical trials is challenging because product risks (e.g., risk of insufficient adverse events reporting or under-enrollment) with uncertain impact on the clinical trial performance dominate the clinical trial risk landscape.

The modeling approach presented in the paper was developed to address the following questions.

a. How to align the contingency budget and product risk^2^?
b. How to allocate a contingency budget optimally across risks, risk categories, and business groups responsible for risk mitigation?
c. How to optimize the risk mitigation strategy for a clinical trial and select an optimal set of mitigation options to reduce clinical trial risk to a certain level of manageability?
d. How can a contingency budget, mitigation strategy, clinical trial risk capacity, appetite, and tolerance be aligned^3^?

The clinical trial risk mitigation complexity (e.g., multiple risks, many mitigation options per risk, risk mitigation effect uncertainty) requires the implementation of the mathematical modeling technique based on a set of interconnected optimization (Risk Optimizer) and simulation (Risk Simulator) models. The Risk Optimizer optimizes the risk mitigation plan and pre-allocates contingency resources, answering questions (a)-(c). The Risk Simulator mimics contingency plan execution and provides recommendations for aligning contingency budget, risk capacity, appetite, and tolerance answering the question (d).

The paper organizes as follows. The first part describes the modeling methodology and its elements. The second part presents the application of the technique to a case study related to a Ph2 clinical trial, including data collection, modeling experiments, and analysis of the results. The paper will conclude with remarks regarding future research.

## Part 1. Modeling methodology

### Risk Optimizer

#### Elements of the model

##### 1. Risk identification and assessment

This paper assumes that each clinical trial risk **i** is characterized by two parameters – impact score (***S***_***i***_) and likelihood score (risk) (***L***_***i***_) in a range (e.g., from 1 to 5 or 1 to 10). The product of these components is a total risk score ***M***_***i***_.

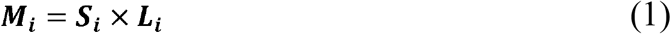

The total risk score is an indicator of risk priority. The higher the total risk score, the more attention should be paid to this risk.

This paper’s risk impact and likelihood scores are subjective parameters scaled from 1 to 5 (Tables 1 and 2).

**Table 1.** Likelihood score Likelihood score **L**_**i**_ scaled from j = (1,5) is a surrogate for an average subjective probability **P**_**i**_ (Li ↔ Pi) or the range of probabilities presented in Table 1. For example, due to assessment subjectivity, a score of five corresponds to “a very high” probability of 90% or probabilities within the 85% - 95% range.

| Likelihood score - L | 1 | 2 | 3 | 4 | 5 |
| --- | --- | --- | --- | --- | --- |
| Rating | Very low | Low | Medium | High | Very high |
| Average, %, P | 10 | 25 | 50 | 75 | 90 |
| Range, % | 5-15 | 15-35 | 35-65 | 65-85 | 85-95 |

**Table 2.** Risk impact. Impact score Si scaled from j = (1,5). It corresponds with qualitative impact assessments from “Very low” to “Very high.”

| Impact - S | 1 | 2 | 3 | 4 | 5 |
| --- | --- | --- | --- | --- | --- |
| Rating | Very low | Low | Medium | High | Very high |

The total risk score (1) for i-th risk can be interpreted as a mathematical expectation (average) of impact ***S***_***i***_. If an i-th risk eventuates, then the impact equals to ***S***_***i***_ (2). If not, the impact ***S***_***i***_ = 0. Therefore, the total risk score ***M***_***i***_ can be presented as

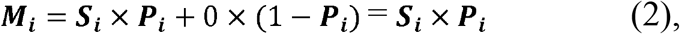

or based on the conversion presented in Table 1 (***L***_***i***_ ↔ ***P***_***i***_)

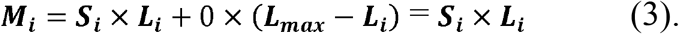

The total risk score ***M***_***i***_ can be interpreted as the average impact of a risk.

Traditionally, risk scores are presented in a risk matrix or “heat map” (Figure 1).

**Figure 1.**
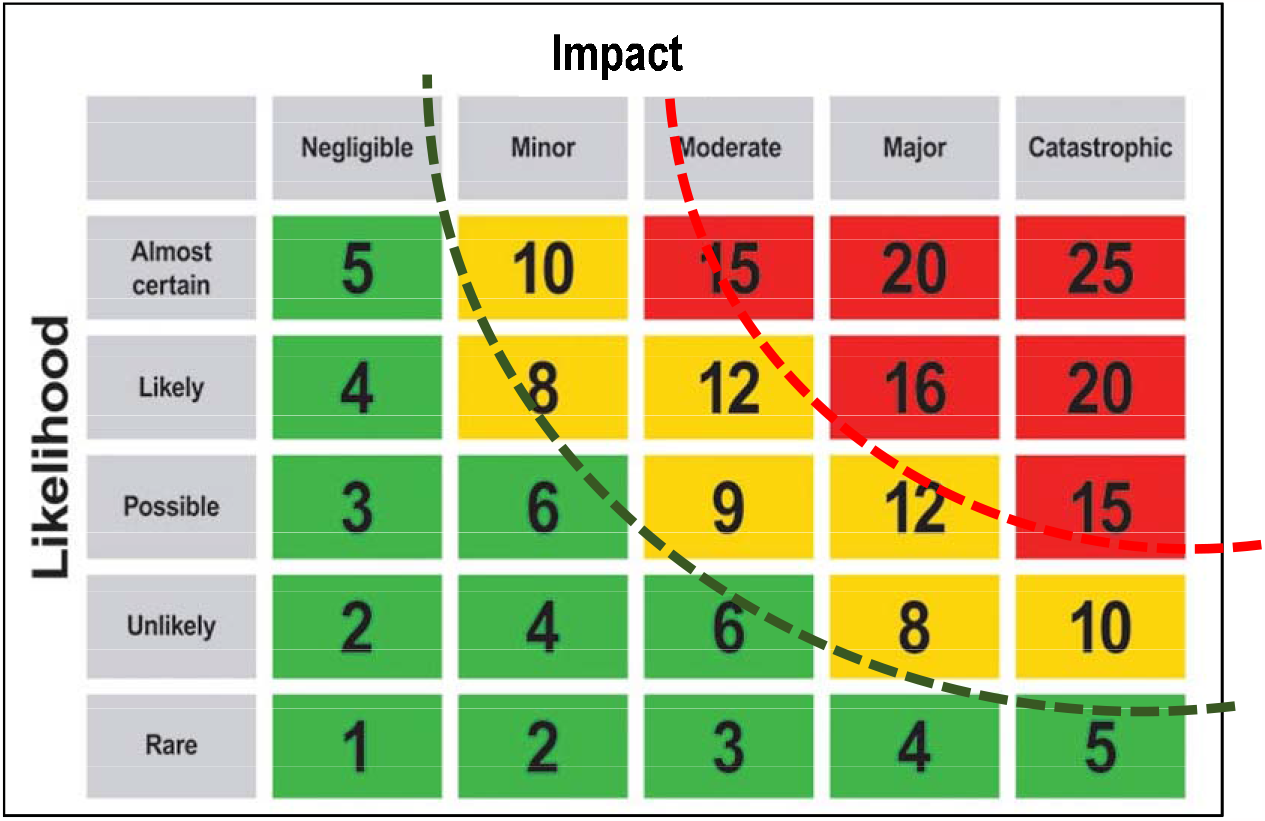
Risk score matrix. If the total risk score for a particular risk is high, the risk falls in the “red” area of a heat map; if medium – in the “yellow” area; and if low – in the “green” area.

The advantages and disadvantages of risk matrices are discussed in [20].

The “heat map” approach could be used to characterize the “riskiness” of the entire clinical trial. The corresponding metric may be presented as a “Total Normalized Risk Score (TNRS).”

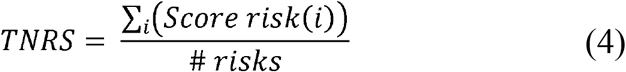

TNRS defined as the total risk score for high and medium risks selected for the analysis divided by the number of analyzed risks.

If TNRS is in the “red” or “yellow” zone at the risk assessment stage, then a mitigation plan for these risks has to be developed. If TNRS is in the “green” zone after developing a mitigation plan, then the mitigation plan could be satisfactory. Zones’ boundaries are arbitrary.

The proposed modeling technique aims to develop a risk mitigation plan for TNRS to be in the “green” zone, depending on a contingency budget.

TNRS has some limitations. For example, although TNRS could be in the “green” zone, some risks could still be in the “red” or “yellow” zone due to limited contingency resources. It means that informal risk analysis should supplement modeling.

TNRS could be a benchmark to help rank clinical trials based on their “riskiness” before and after the mitigation.

##### 2. Risk mitigation

A risk mitigation plan has to answer the critical question in risk planning – what to do if a risk event occurs. There are many options, such as (a) to leave it as is and face unfavorable consequences; (b) to eliminate risk by allocating significant contingency resources, or (c) to find the right balance between the degree of risk mitigation, contingency budget, risk appetite^4^, risk capacity^5^, and risk tolerance^6^, assuming that any mitigation plan is associated with residual risks^7^.

Both under- and over-allocation of contingency resources are suboptimal. Under-allocation of the contingency budget could jeopardize clinical trial execution. Over-allocation of a contingency budget, or even incorporating additional resources into a core clinical trial budget to eliminate some risks (assuming that probability is almost 100% which may not be the case), could increase a clinical trial budget.

#### Mitigation costs

The contingency budget comprises mitigation costs required to mitigate each risk upon its occurrence. They are assigned to each risk mitigation option. The presented technique allows the incorporation of multiple mutually exclusive risk mitigation options associated with a single risk.

Deeper risk mitigation corresponds with higher mitigation costs. Training, monitoring visits, or backup sites are typical risk mitigation options. Each risk is associated with the probability of occurrence. Therefore, the contingency costs should be multiplied by the probability of occurrence (Table 1) for each risk (5), including reduced risks after the mitigation.

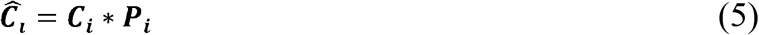

where 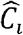- risk-adjusted (average) risk mitigation costs; ***C***_***i***_ – mitigation costs, and ***P***_***i***_ – the probability of occurrence for i-th risk.

Several empirical approaches are known to address contingency planning in clinical trial budgeting.

a. The first approach is based on identifying the risks with the highest risk score and allocating contingency resources to eliminate these risks.
b. The second approach assumes contingency resource allocation for the preselected risks.

Techniques (a) and (b) do not guarantee optimal solutions. Therefore, with increased clinical R&D budgetary constraints and complexities of clinical trials, the need for a method that accurately aligns risk, contingency resources, risk appetite, and risk tolerance cannot be underestimated.

#### Contingency planning methodology for a clinical trial

The effective risk planning process during clinical trial budgeting needs to answer the following questions:

1. How to align the contingency budget with the clinical trial risk landscape?
2. What contingency budget should be allocated to reduce overall clinical trial risk to a certain level, e.g., to reduce TNRS to the green zone (Figure 1))?
3. Should the contingency budget be allocated to eliminate (1) all risks, (2) the highest risks, or (3) reduce TNRS to a required level by optimizing it?
4. How to align risk mitigation, contingency budget, and risk tolerance?

##### 3. The model

The model was developed to optimize the risk mitigation plan. A risk score is calculated for each identified risk from the risk registry using the formula (1). It is assumed that low-score risks are accepted. For high and medium risks, one or multiple risk mitigation options need to be identified along with resources to reduce them in case of risk eventuation. The corresponding residual risk score^8^ is also calculated for each mitigation option.

The model selects an optimal risk mitigation plan allocating contingency budget across risks minimizing TNRS for the entire portfolio of clinical trial risks or minimizing the contingency budget aiming to “move” TNRS into the “green” zone of the risk matrix (Figure 1). It also allows the optimal allocation of limited contingency resources across different business groups and risk categories.

The model is formulated as a Mixed Integer Programming (MIP) model [21] where some variables are binary (X= 0, 1). MIP models find the globally optimal value of a function for a certain number of variables, given a set of constraints on these variables (equalities or inequalities).

The model includes four components:

##### A. Decision variables

X_i_ = (0 or 1). If X_i_ = 0, the i-th risk/risk mitigation option is not selected. If X_i_ = 1, the i-th risk/risk mitigation option is selected. Their value will be defined automatically during the model execution.

##### B. Parameters

All model parameters are estimates.

▪ For each risk and risk mitigation option - the risk score is calculated using (1)
▪ Risk-adjusted mitigation cost is calculated using (5) for each risk mitigation option.
▪ The average contingency budget.

##### C. Constraints and rules

▪ The average contingency budget should exceed or be equal to the total average mitigation costs for selected risk mitigation options
▪ Business rules (e.g., forced elimination of risks, forced reduction of highest risks’ likelihoods, etc.)
▪ Others

##### D. Criteria

Criteria may include:

1. Minimum TNRS for selected risks
2. Minimum quadratic TNRS deviation from the TNRS target.
3. Minimum contingency budget
4. Others (e.g., multiple criteria could be used)

The developed technique was applied to the risk planning of a Ph2 clinical trial.

## Part 2. Case study

### 1. Risks identification and assessment

For a clinical trial, 80 risks^9^ across ten risk categories were identified. The risks were separated into two categories – low and medium/high. The number of low risks = 55, number of high/medium risks = 25. Risks allocation across risk categories is presented in Figure 2. Risk categories, risks, and allocation across risk categories are unique for each clinical trial.

**Figure 2.**
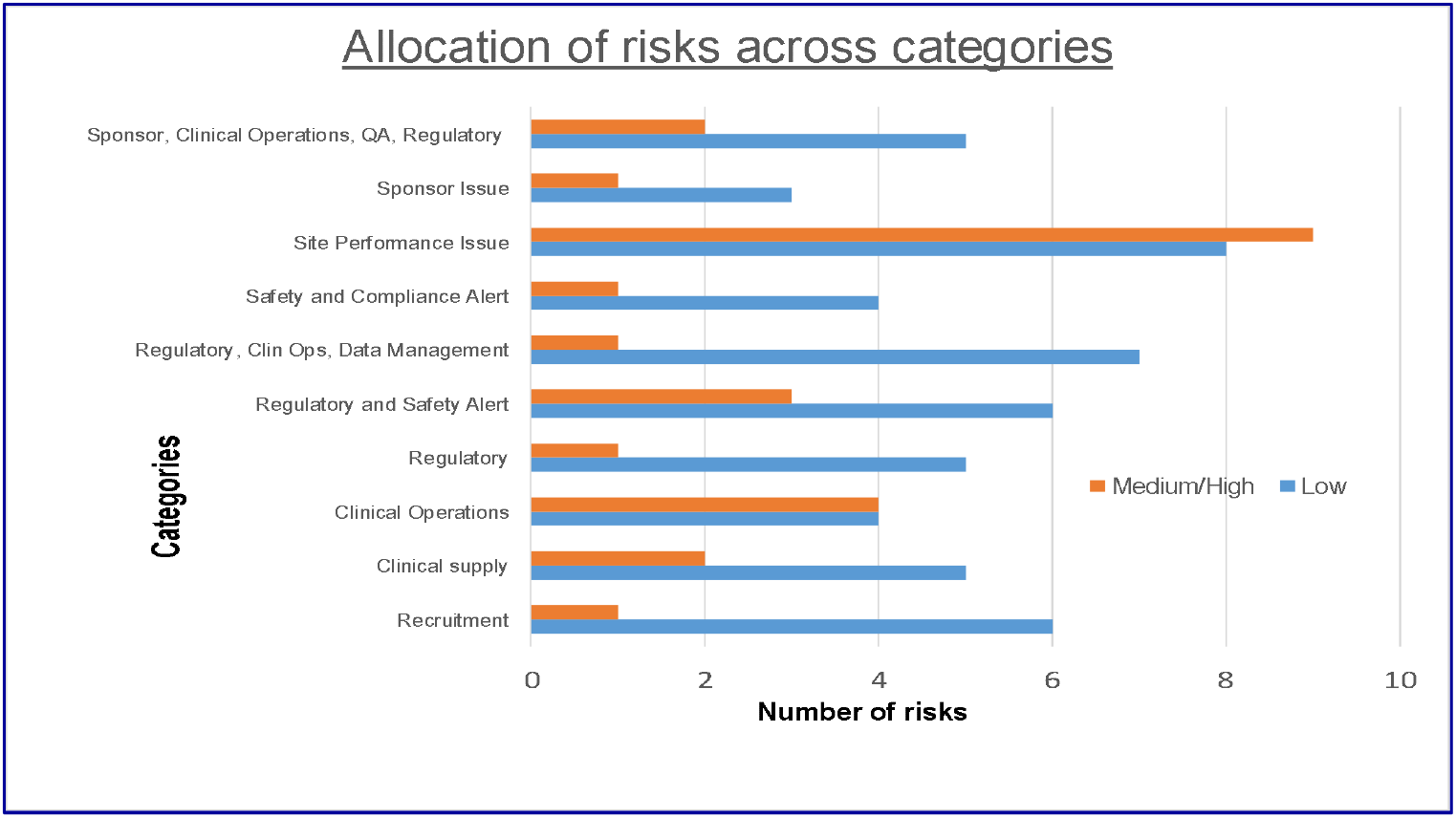
Allocation of low and medium/high risks across risk categories.

Low risks may not be as harmful as medium/high risks for clinical trial performance. Therefore, they may be accepted. Medium/high risks are mitigated (mitigation costs are assigned to each mitigation option, see Table 3) in the case study to minimize their impact on clinical trial performance. Mitigation cost assignment to each risk and corresponding mitigation options are associated with their high detectability. Therefore it is assumed that the level of detectability across mitigated risks is the same, and it does not affect the modeling results.

**Table 3.** Fragment of the risk registry with risk mitigation options and risk mitigation costs (one mitigation option per risk).

| Risk # | Category | Mitigation option | Description of risk and risk mitigation | Likelihood score | Impact score | Total risk score | Mitigation cost, \$ | Probability | Risk-adjusted mitigation costs |
| --- | --- | --- | --- | --- | --- | --- | --- | --- | --- |
| 1 | Recruitment |  | Recruitment is not completed as planned |  |  |  |  |  |  |
|  |  | 1.1 | 10% fewer patients are recruited during the first three months of the study. **No action** | 5 | 4 | 20 | 0 | 0.9 | 0 |
|  |  | 1.2 | 22 sites in two new countries | 1 | 4 | 4 | 720,000 | 0.1 | 72,000 |
| 2 | Clinical Operations |  | The study cannot be completed on time due to delays in contracting with sites |  |  |  |  |  |  |
|  |  | 2.1 | A one-month delay at 25% of sites. **No action** | 4 | 4 | 16 | 0 | 0.75 | 0 |
|  |  | 2.2 | Add 100h required resources | 2 | 4 | 8 | 10000 | 0.25 | 2500 |

### Risk mitigation planning

The proposed methodology focuses on deriving an optimal risk mitigation strategy for a clinical trial’s entire portfolio of risks.

### Input data

The approach is illustrated in Tables 3 and 4. In Table 3, it is assumed that each risk has two options: (1) no mitigation and (2) to mitigate risk to minimize its score. For example, risk #1 is related to a lower-than-expected number of patients (10% fewer patients are enrolled during the first three months of the study). Both impact and probability of occurrence for this risk are high (4 and 5). Overall risk score = 20. The mitigation option for risk #1 is to set up 22 new sites in two new countries. The mitigation cost is 720K. The pre-allocation of 22 backup sites will reduce risk#1 likelihood to one and the residual risk score to four. According to Table 2, the probability of risk #1 was reduced from 85%-95% to about 5% - 15% (average -10%, Table 3). Therefore, in contingency planning, the probability-adjusted cost for risk #1, according to (5), is calculated as 720K*0.1=72K.

**Table 4.** Fragment of the risk registry with risk mitigation options and risk mitigation costs (multiple mitigation options per risk).

| Risk # | Category | Mitigation option | Description of risk and risk mitigation | Likelihood score | Impact score | Total risk score | Mitigation cost, \$ | Probability | Risk-adjusted mitigation costs |
| --- | --- | --- | --- | --- | --- | --- | --- | --- | --- |
| 1 | Recruitment |  | Recruitment is not completed as planned |  |  |  |  |  |  |
|  |  | 1 | 10% fewer patients are recruited during the first three months of the study. **No action** | 5 | 4 | 20 | 0 | 0.9 | 0 |
|  |  | 1.1 | 3 new sites in 5 current countries | 4 | 4 | 16 | 40,000 | 0.75 | 30,000 |
|  |  | 1.2 | 4 new sites in 2 new countries | 3 | 4 | 12 | 90,000 | 0.5 | 45,000 |
|  |  | 1.3 | 10 new sites in 5 current countries | 2 | 4 | 8 | 200,000 | 0.25 | 50,000 |
|  |  | 1.4 | 22 sites in two new countries | 1 | 4 | 4 | 720,000 | 0.1 | 72,000 |
| 2 | Clinical Operations | 2 | The study cannot be completed on time due to delays in contracting with sites |  |  |  |  |  |  |
|  |  | 2.1 | A one-month delay at 25% of sites. **No action** | 4 | 4 | 16 | 0 | 0.75 | 0 |
|  |  | 2.2 | Add 100h required resources | 2 | 4 | 8 | 10000 | 0.25 | 2500 |

2^**25**^ = ∼33M+ options – to mitigate risk or not – could be potentially considered to develop the most effective risk mitigation plan for identified 25 medium/high risks. It is time-consuming to analyze even a small part of them. If a contingency budget is limited, how to decide which risk to mitigate and which is not?

The optimization model allows the incorporation of multiple mutually exclusive risk mitigation options per risk, including “no action,” as presented in Table 4. In this example, risk #1 (the risk that recruitment is not completed as planned) has five mutually exclusive risk mitigation options related to the potential activation of new sites in current and new countries with corresponding mitigation costs.

Three models are considered to select the most effective selection algorithm – (A) – the model based on risk sorting; (B) – an optimal risk mitigation algorithm with one mitigation option per risk; and (C) - an optimal risk mitigation algorithm with multiple mutually exclusive mitigation options per risk providing additional flexibility in decision-making.

#### A. Sorting algorithm

Many practitioners use the sorting or prioritization algorithm to sort risks according to their total risk score. Related cumulative risk-adjusted mitigation costs are added until they exceed the contingency budget. Selected risks allocated resources are included in a risk mitigation plan.

#### B. An optimal risk mitigation algorithm-one mitigation option per risk

The risk mitigation plan based on risk sorting can be improved using the optimization model described above.

#### C. An optimal risk mitigation algorithm-multiple mitigation options per risk

##### Modeling results. Baseline scenario

According to Figure 3, the clinical trial TNRS would be about 30% lower if the optimization model (B) had been used compared to the sorting (A) and about 35% for model (C) for a contingency budget of 300K. Both algorithms generate the same results if the contingency budget is zero (no mitigation) or exceeds 650K (large contingency budget - mitigation of all risks).

**Figure 3.**
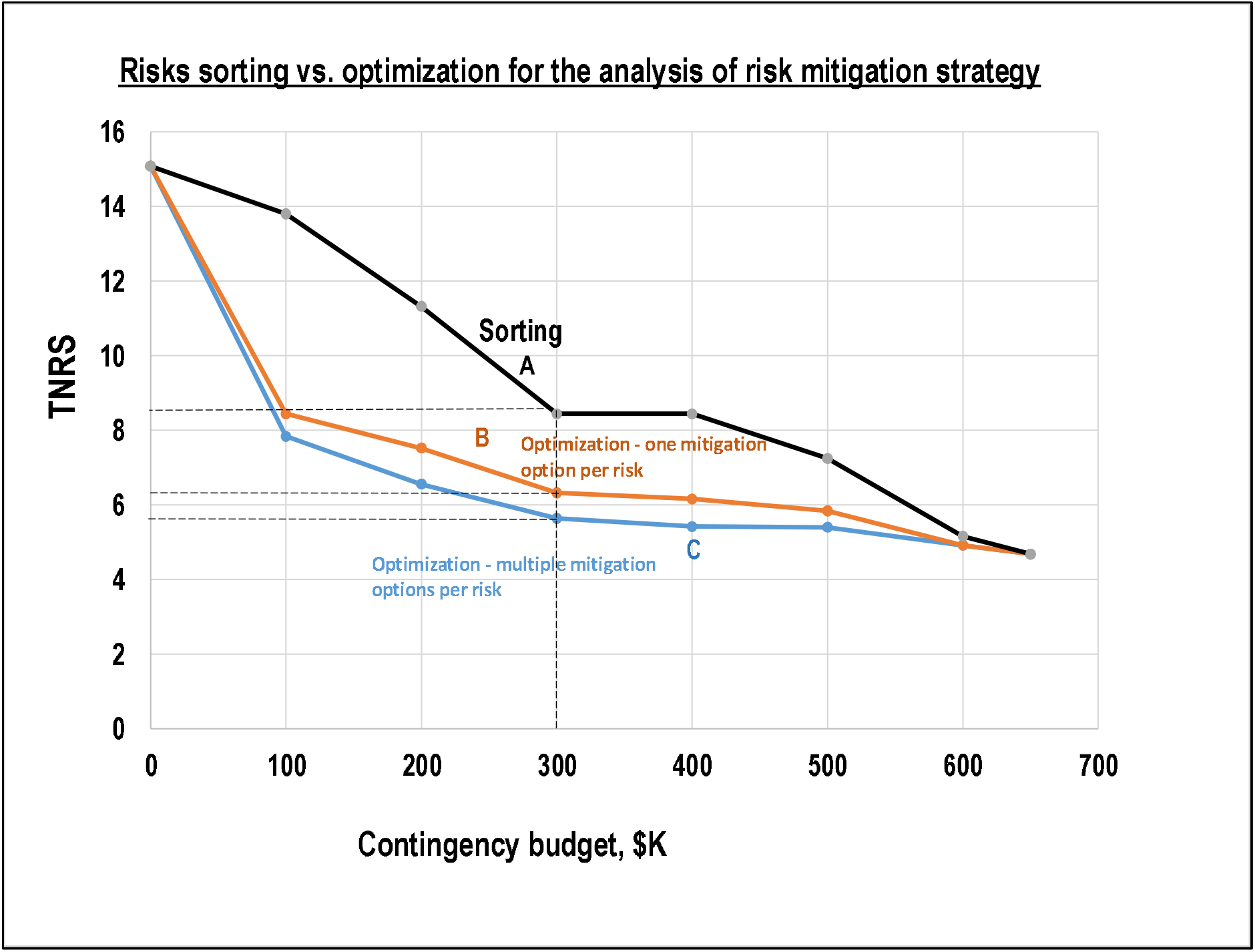
Comparison of risk mitigation planning algorithms. A – sorting; B – optimization (one mitigation option per risk); C optimization (multiple mitigation options per risk) for different contingency budgets from zero to 650K.

Table A (Appendix 1) presents the optimal risk mitigation plan for the baseline scenario.

##### Contingency resources allocation

The model generates an optimal allocation of contingency budget across risk categories (see Figure 1 for a list of risk categories), as shown in Figure 4.

**Figure 4.**
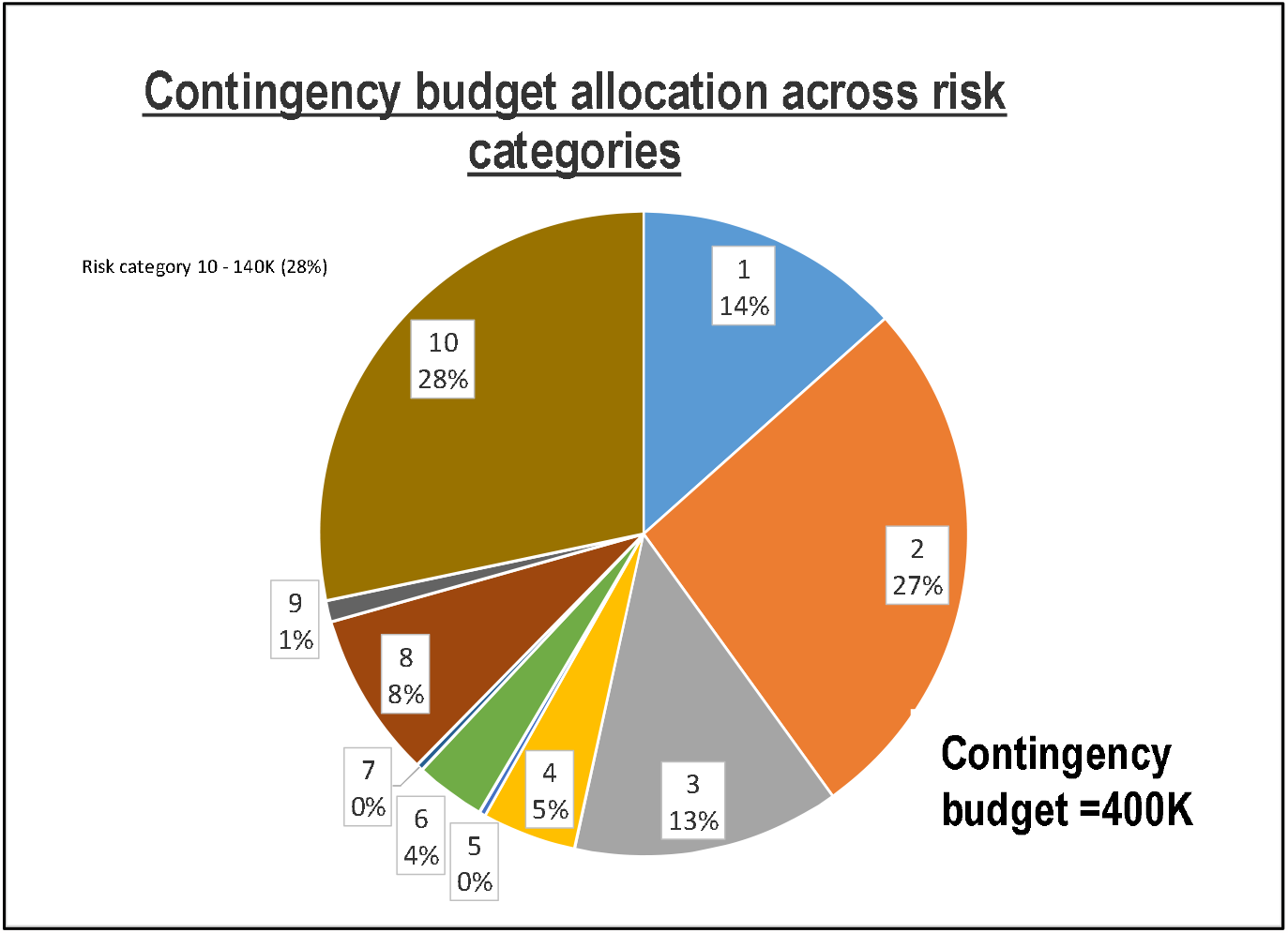
Contingency budget allocation across risk categories. 28% of a contingency budget is allocated for the risk category #10

###### b. Model analysis. What-if scenarios

Two “what-if” scenarios were generated and compared with the baseline scenario.

###### 1. Forced selection of mitigation option #25.2 of risk#25 for contingency budget = 400K. Risks interdependence

In the baseline scenario, risk #25, related to the high risk of stock-out in clinical supplies of a clinical trial, was not mitigated due to a limited contingency budget (Table 5). Risks #23 and #24 were mitigated. The model selected options #23.1 and #24.1 (plus sign). The decision was made to force the mitigation of risk #25 to minimize the risk of clinical supply stock-out with maximum overage. It means allocating 225K to minimize the risk score (forced selection of option #25.2). If option #25.2 for risk #25 is forced to be selected, then risks #23 and #24 cannot be mitigated (Table 6). The proposed risk plan is riskier (TNRS = 5.84 vs. 5.12 in the baseline scenario for the 400K contingency budget).

**Table 5.** Fragment of baseline scenario with selected mitigation options

| # | Risks | Baseline scenario | Mitigation costs, K |
| --- | --- | --- | --- |
| 23 | Batch failure rate is high |  | 0 |
| 23.1 | Backup – CMO #1 | + | 100 |
| 23.2 | Backup – CMO #2 |  | 150 |
| 24 | Slow drug distribution and resupply process |  | 0 |
| 24.1 | Faster delivery. Backup carrier | + | 125 |
| 24.2 | Faster delivery options + Additional depots |  | 175 |
| 25 | High risk of stock-out | + | 0 |
| 25.1 | Increase overage by 5% |  | 200 |
| 25.2 | Increase overage by 10% |  | 225 |

**Table 6.** Forced selection of option 2 for risk #25 vs. baseline scenario.

| Risk | Forced risk #25 reduction | Baseline scenario |
| --- | --- | --- |
| 23 | 1 | 0 |
| 23.1 | 0 | 1 |
| 23.2 | 0 | 0 |
| 24 | 1 | 0 |
| 24.1 | 0 | 1 |
| 24.2 | 0 | 0 |
| 25 | 0 | 1 |
| 25.1 | 0 | 0 |
| 25.2 | 1 | 0 |
| **TNRS** | **5.84** | **5.12** |

###### 2. Forced mitigation of high risks

The model (with additional rules) can force the selection of mitigation options to reduce their highest likelihood; for example, it can force the reduction of the likelihood of (≥4).

Figure 5 illustrates the dynamics of TNRS for three scenarios: (1) - Baseline scenario; (2) - Forced mitigation of risk #25 (forced selection of option #25.2); and (3) – forced mitigation of risks with a high likelihood (≥4) for different values of a contingency budget.

**Figure 5.**
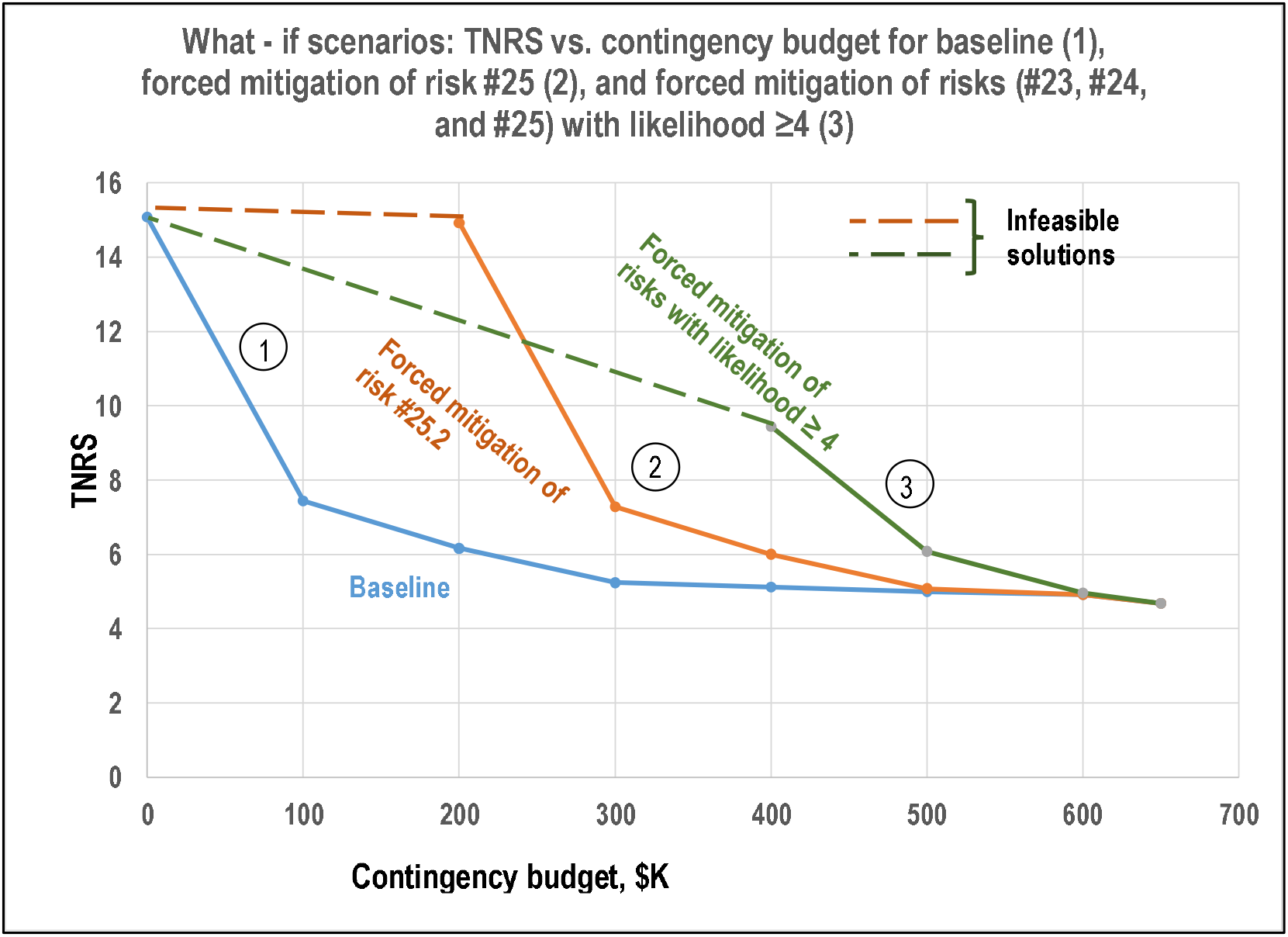
What-if scenarios. (1) - Baseline scenario; (2) - Forced mitigation of risk #25 (forced selection of option #25.2); and (3) – forced mitigation of risks with a high likelihood (≥4).

Modeling experiments indicate that the contingency budget for scenario #2 could generate feasible scenarios (plausible mitigation strategies) if it exceeds 200K to meet the rule to force mitigation of risk #25. The contingency budget should exceed 400K to meet the likelihood reduction rule (scenario #3). Otherwise, the model will generate infeasible solutions (dotted lines), and the mitigation strategy will not be acceptable.

### Alignment of contingency budget, risk tolerance, and risk appetite

The optimal contingency planning model described above generates an optimal risk mitigation plan using “averaged” data such as the optimization criteria, risk mitigation costs, and contingency budget. It addressed many essential questions, such as optimal risk mitigation strategy selection and contingency budget allocation. However, the model cannot answer how to incorporate risk appetite, capacity, and tolerance into the contingency budgeting process.

### Risk appetite, capacity, and tolerance

are associated with uncertainty due to different stakeholders’ expectations. Therefore, it seems natural to define risk tolerance as the probability that a contingency budget will be sufficient to implement a mitigation plan, assuming residual impact and probability for each risk. A lower risk tolerance requires a higher contingency budget and vice versa.

For the risk appetite, capacity, and tolerance assessment, the portfolio of mitigated risks was simulated using binary distribution to analyze risk tolerance (Figure 6).

**Figure 6.**
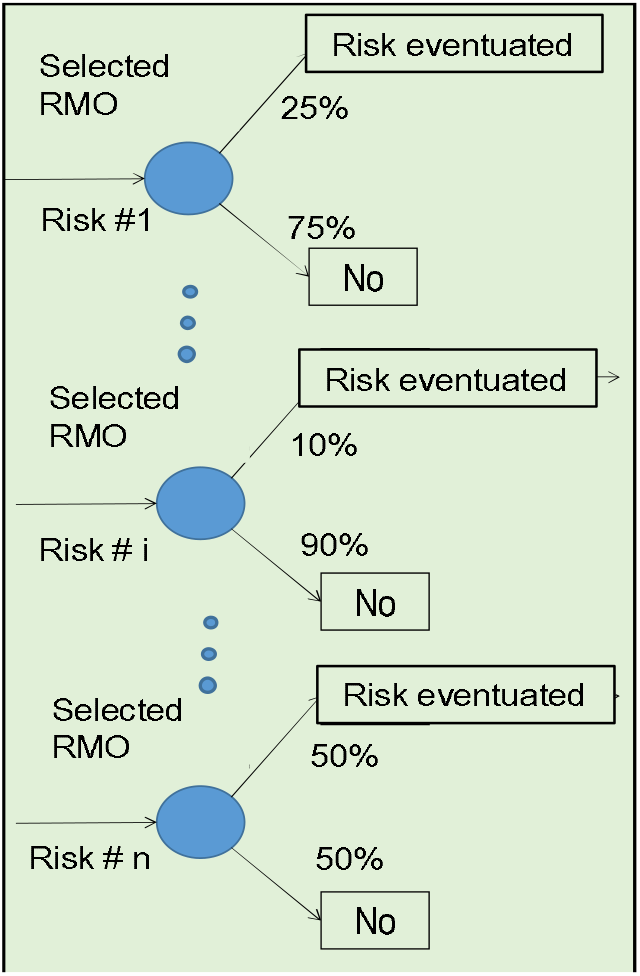
Monte-Carlo simulation of selected risk mitigation strategy for the portfolio of clinical trial risks. RMO – Risk Mitigation Option

### Simulation results

A contingency budget = 300K (Figure 7) corresponds to 50% risk tolerance. It means that if a contingency budget equals 300K, there is only a 50% chance that the mitigation strategy will be implemented. If a contingency budget equals 600K, there is an 80% chance that the mitigation strategy will be implemented (20% risk tolerance). Zero risk tolerance (∼100% that the mitigation strategy will be implemented) requires a ∼1.43M contingency budget. Risk capacity is defined as 90% (10% tolerance limit) and fits with a ∼1M contingency budget. Risk appetite (20%) could correspond to a 600K contingency budget.

**Figure 7.**
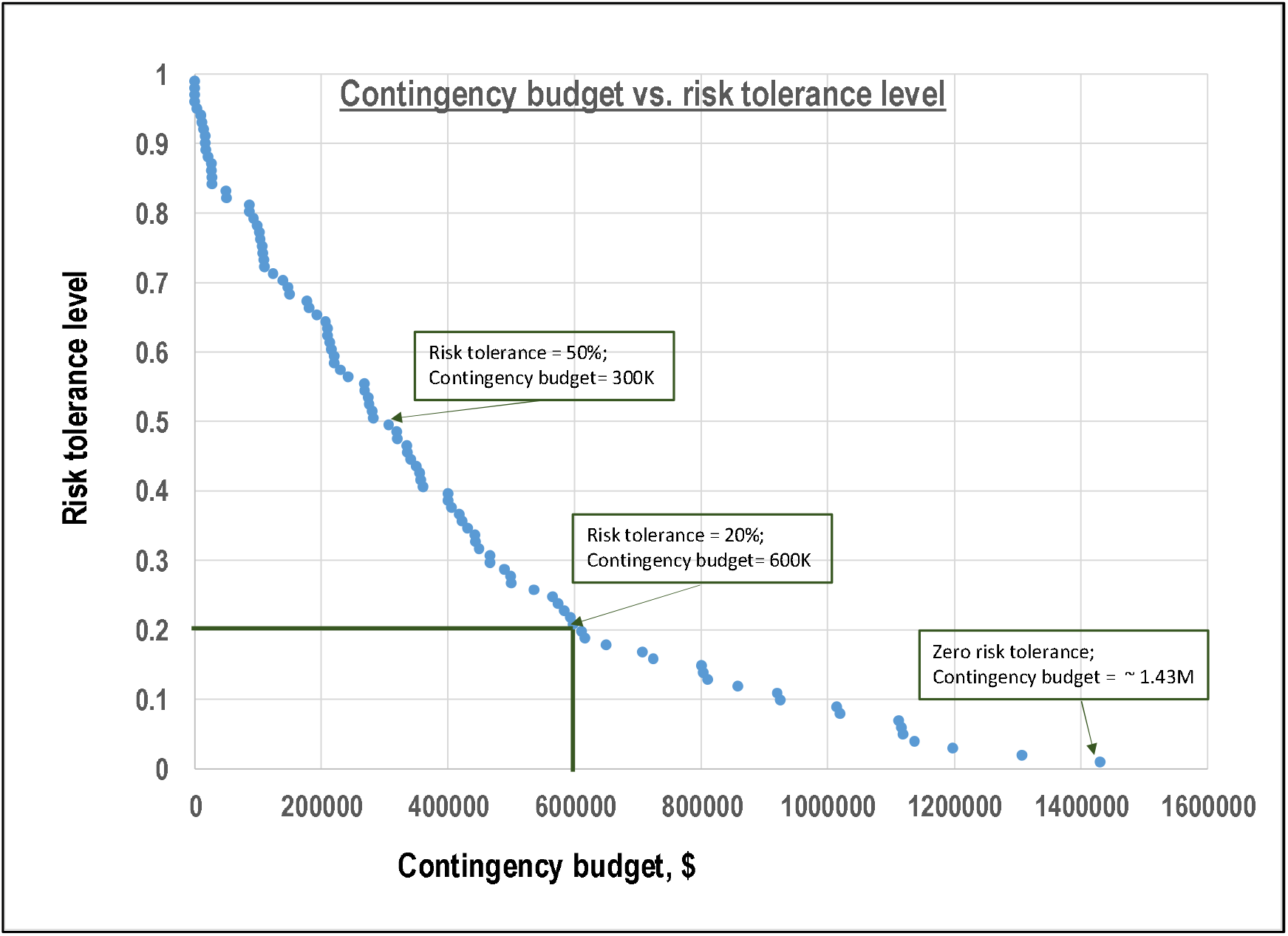
Simulation for a portfolio of risks – empirical probability distribution The horizontal axis is the contingency budget, and the vertical is the risk tolerance level: the lower the risk tolerance, the higher the contingency budget.

Due to resource limitations, small companies may tolerate a higher risk than big ones.

### Future research

Future research will focus on deriving the optimal values of QTL/KRI and secondary limits aligned with a clinical trial risk landscape and mitigation resources based on the optimal allocation of the contingency budget.

### Conclusion

- Effective contingency planning in clinical trial budgeting could substantially reduce risk and save money
- A methodology for optimal risk mitigation planning was developed and validated. It minimizes total normalized risk score (TNRS) under a limited budget and utilizes multiple risk mitigation options per risk
- Modeling experiments demonstrated the effectiveness and efficiency of the model

The author thanks Andor Kanka and Dr. Beata Balati (Balati CRO, Hungary) for sharing data and constructive discussions and anonymous reviewers for valuable comments.

## Data Availability

All data produced in the present work are contained in the manuscript

## Footnotes

1 Quantitative risk analysis can be used only if data is available to measure and predict a risk event’s probability and impact (e.g., project delay, loss of revenue, etc.). It typically involves numerical analyses of the effects of risks, such as time and cost.

2 For simplicity, the term “risk” will be used in the paper instead of “product risk.”

3 In the paper, risk tolerance is defined as the probability that a selected mitigation strategy will be accomplished

4 **Risk appetite** is the amount of risk a clinical trial planner desires to take

5 **Risk capacity** is the maximum amount of risk that could be taken.

6 **Risk tolerance** is the amount of risk a clinical trial planner is comfortable taking or the degree of uncertainty the planner can handle.

7 All risks in the case study will be monitored.

8 If mitigation resources are assigned, it is assumed that the residual risk still exists.

9 Assuming they are independent

## References

1. Ghone A. Risk Assessment In Clinical Trials – Well Begun Is Half Done. https://www.clinicalleader.com/doc/risk-assessment-in-clinical-trials-well-begun-is-half-done-0001

2. Aven T. Risk assessment and risk management: Review of recent advances on their foundation. European Journal of Operational Research 253 (2016) 1–13

3. U.S. Department of Health and Human Services Food and Drug Administration, E6 (R2) Good Clinical Practice: Integrated Addendum to ICH E6(R1) Guidance for Industry ICH E6. https://www.fda.gov/downloads/Drugs/Guidances/UCM464506.pdf (2018).

4. Della-Coletta, A., Katz, T., Kupka, K. et al. Toolkit for ICH E6 (R2) Quality Risk Management for Small to Medium Size Companies. Ther Innov Regul Sci 54, 900–921 (2020).

5. Liebig H, Hastings R. Reducing Risk through Mitigation Strategies. Applied Clinical Trials, 2009, August.

6. Bacchieri A, Rossi A, Morelli P. Risk and mitigation actions for clinical trials during COVID-19 pandemic (RiMiCOPa). Contemporary Clinical Trials Communications 20 (2020).

7. Knirsch C, Chappell P, Alvir J, Alemayehu D. Risk Assessment and Mitigation. Applied Clinical Trials, 2012, Volume 21, Issue 4

8. Del Medico A, Trad M, Khinda S, Moody B, Hughes L, Vanbelle C, Tanjga O. Mitigating Risk in Implementing Multi-Regional Trials in M.S. Applied Clinical Trials-02-01-2018, Volume 27, Issue 2.

9. Bhagat S, Kapatkar V.K, Mourya M, Roy S, Jha S, Reddy R, Kadhe G, Mane A, Sawant S. Potential Risks and Mitigation Strategies Before the Conduct of a Clinical Trial: An Industry Perspective Reviews on Recent Clinical Trials, 2016, 11, 47–55

10. Bowers J, Khorakian A. Integrating risk management in the innovation project. European Journal of Innovation Management 2014 Vol. 17 No. 1, pp. 25–40

11. Mordecai Y. Project risk optimization based on strategy-proof risk assessment. M.S. Industrial Engineering thesis. Tel-Aviv University. May 2010

12. Guan X, Servranckx T, Vanhoucke M. An analytical model for budget allocation in risk prevention and risk protection. Computers & Industrial Engineering 161 (2021)

13. Micán C, Fernandes G, Araújo M. Project portfolio risk management: a structured literature review with future directions for research International Journal of Information Systems and Project Management, Vol. 8, No. 3, 2020, pp. 67–84.

14. Ben-David, I., & Raz, T. (2001). An integrated approach for risk response development in project planning. Journal of the Operational Research Society, 52(1), 14–25.

15. Chapman C. Project risk analysis and management - PRAM is the generic process. International Journal of Project Management, Volume 15, Issue 5, October 1997, Pp. 273–281.

16. Sato T, Hirao M. Optimum budget allocation method for projects with critical risks. International Journal of Project Management, Volume 31, Issue 1, January 2013, pp. 126–135.

17. Cagno E, Caron F, Mancini M. Dynamic analysis of project risk. Int. J. Risk Assessment and Management, 2008, Vol. 10

18. Thaheem M, De Marco A, Narbaev T. Selecting Appropriate Risk Response Strategies Considering Utility Function and Budget Constraints: A Case Study of a Construction Company. Buildings 2022, 12, 98. Pp.1–19

19. M. Grabowski M, Roberts K. Risk Mitigation in Large-Scale Systems: Lessons from high-reliability organizations. California management review vol 39, no. 4 summer 1997

20. Cox T. What’s Wrong with Risk Matrices? Risk Analysis, Vol. 28, No. 2, pp. 497–512.

21. Dorfman, R., Samuelson, P. A., & Solow, R. M. (1986). Linear programming and economic analysis. Revised edition. Mineola, NY: Dover Books on Computer Science.

