## Supplemental Table 1 for "Efficient risk mitigation planning for a clinical trial" (from another version of this preprint: version 1, 2022-11-03)

| Risk mitigation plan  Appendix 1. Table A. Model generated risk mitigation plan for a clinical trial (example data) | | | **Severity  score (S)** | **Prob. Of  occurrence (L)** | **Total  score (SxL)** | **Mitigation  option selected** |
| --- | --- | --- | --- | --- | --- | --- |
| Risk | 1 | **Risk of low enrollment** | | | | |
| Mitigations | 1.1 | 10% fewer subjects are enrolled during the first three months of the study. **No action** | 4 | 5 | 20 | 0 |
|  | 1.2 | Three new sites are opened in 5 currently involved countries | 4 | 4 | 16 | 0 |
|  | 1.3 | Four new sites in two new countries | 4 | 3 | 12 | 0 |
|  | 1.4 | Ten new sites are in 5 currently involved countries | 3 | 3 | 9 | 0 |
|  | **1.5** | **Ten new sites in two new countries** | **3** | **2** | **6** | **1** |
|  | 1.6 | 22 new sites in 5 currently involved countries | 2 | 2 | 4 | 0 |
|  | 1.7 | 22 new sites in two new countries | 1 | 2 | 2 | 0 |
| Risk | **2** | **The study cannot be completed on time due to a delay in contracting with the sites** | | | | |
| Mitigations | 2.1 | There is one month delay of 25% sites. **No action** | 4 | 4 | 16 | 0 |
|  | 2.2 | Add CRA resources 100h CRAs | 1 | 2 | 2 | 1 |
| Risk | **3** | **The dropout rate during the study was higher than expected by the end of the recruitment period. The following risk indicators and thresholds were defined:** | | | | |
| Mitigations | 3.1 | The dropout rate measure up to 2% when 90% of the planned subjects are randomized - **no action** | 4 | 4 | 16 | 0 |
|  | 3.2 | The recruitment period is extended, and 8% more subjects are randomized by one month | 1 | 2 | 2 | 1 |
| **Risk** | **4** | **Delay in obtaining regulatory and ethics committee approvals. There are two months delay in study start-up; the planned milestones are expenses by two months until regulatory approval is obtained and sites activated** | | | | |
| Mitigations | 4.1 | Two months delay accepted, **no action** | 4 | 5 | 20 | 0 |
|  | 4.2 | Three new sites open in one country - 1-month delay (add risk #1) | 3 | 3 | 9 | 0 |
|  | 4.3 | Five new sites open in two countries - no delay (add risk #1) | 1 | 1 | 1 | 1 |
| **Risk** | **5** | **There is a 1-month delay in data cleaning and database lock, and therefore, the study may be completed one month later than planned** | | | | |
| Mitigations | 5.1 | Accept a 1-month delay, **no action** | 4 | 5 | 20 | 0 |
|  | 5.2 | More data manager and clinical monitor time is allocated  for data cleaning and DB lock (limited workload) (50h) | 4 | 3 | 12 | 0 |
|  | 5.3 | More data manager and clinical monitor time is allocated  for data cleaning and DB lock (limited workload) (80h) | 3 | 3 | 9 | 0 |
|  | 5.4 | More data manager and clinical monitor time is allocated  for data cleaning and DB lock (limited workload) (100h) | 1 | 2 | 2 | 1 |
|  | 5.5 | More data manager and clinical monitor time is allocated  for data cleaning and DB lock (limited workload) (130h) | 1 | 2 | 2 | 0 |
| **Risk** | **6** | **High site personnel turnover at a number of sites** | | | | |
| Mitigations | 6.1 | There is a significant delay in data entry, personal changes, the volume of workload at ten sites, and **no action** | 3 | 4 | 12 | 0 |
|  | 6.2 | Ten site retraining visits are necessary, eight h each | 2 | 2 | 4 | 1 |
| **Risk** | **7** | **Delayed query resolution at several sites** | | | | |
| Mitigations | 7.1 | There is a significant delay in query resolution at 20 sites - no action | 5 | 4 | 20 | 0 |
|  | 7.2 | 80 hours of extra time in site management is necessary | 2 | 1 | 2 | 1 |
| **Risk** | 8 | **Data trend abnormalities were detected at some sites** | | | | |
| Mitigations | 8.1 | Data trend abnormalities detected at ten sites - no action | 4 | 5 | 20 | 0 |
|  | 8.2 | Five extra monitoring visits/sites are necessary | 1 | 1 | 1 | 1 |
| **Risk** | 9 | **Repeat site errors indicate the inexperience of site personnel  at some sites** | | | | |
| Mitigations | 9.1 | Inexperienced site personnel at ten sites - no action | 4 | 3 | 12 | 0 |
|  | 9.2 | Ten retraining visits are necessary (one visit/site) | 1 | 1 | 1 | 1 |
| **Risk** | 10 | **Over enrollment (50% more subjects enrolled than planned) at some sites** | | | | |
| Mitigations | 10.1 | Over enrollment at ten sites - no action. Clinical supplies could be disrupted. | 4 | 3 | 12 | 0 |
|  | 10.2 | Clinical supply plans corrections. Visits rescheduling. Stayed with current resources | 1 | 1 | 1 | 1 |
| **Risk** | 11 | **Delayed data entry at sites** | | | | |
| Mitigations | 11.1 | data entry delay at 20 sites - no action | 4 | 3 | 12 | 0 |
|  | 11.2 | 40 hours of extra site management time is necessary | 3 | 2 | 6 | 0 |
|  | 11.3 | 60 hours of extra site management time is necessary | 3 | 2 | 6 | 0 |
|  | 11.4 | 70 hours of extra site management time is necessary | 1 | 1 | 1 | 1 |
| **Risk** | **12** | **Eligibility (inclusion/exclusion) criteria were not met for some sites** | | | | |
| Mitigations | 12.1 | Eligibility criteria violation detected at ten sites - no action | 4 | 3 | 12 | 0 |
|  | 12.2 | Ten retraining visits and 20 more monitoring visits are necessary at ten sites | 1 | 1 | 1 | 1 |
| **Risk** | **13** | **The query rate is significantly higher than the average for some sites** | | | | |
| Mitigations | 13.1 | The query rate is significantly higher at ten sites - no action | 4 | 4 | 16 | 0 |
|  | 13.2 | 25% more monitoring visits are necessary at ten sites | 2 | 2 | 4 | 1 |
| **Risk** | **14** | **The number of protocol violations is higher at some sites** | | | | |
| Mitigations | 14.1 | The number of protocol violations is higher at ten sites - no action | 4 | 4 | 16 | 0 |
|  | 14.2 | Ten retraining visits and 20 more monitoring visits are necessary at ten sites | 1 | 1 | 1 | 1 |
| **Risk** | **15** | **Repeated unreported SAE detected at five sites** | | | | |
| Mitigations | 15.1 | Repeated unreported SAE detected at five sites - no action | 3 | 4 | 12 | 0 |
|  | 15.2 | Ten more monitoring visits are necessary to perform a 100% SDR review at five sites | 2 | 1 | 2 | 1 |
| **Risk** | **16** | **The wrong version of the protocol was used to conduct the study** | | | | |
| Mitigations | 16.1 | The wrong version of the protocol was used to conduct the study at two sites. No action | 3 | 4 | 12 | 0 |
|  | 16.2 | Four more monitoring visits are necessary | 1 | 1 | 1 | 1 |
| **Risk** | **17** | **Informed consent was not properly obtained nor documented at multiple sites** | | | | |
| Mitigations | 17.1 | Informed consent was not properly obtained nor documented at five sites - no action | 3 | 3 | 9 | 0 |
|  | 17.2 | one retraining visit/site is necessary | 2 | 1 | 2 | 1 |
| **Risk** | **18** | **Protocol inconsistencies were recognized after several months from the start of the study** | | | | |
| Mitigations | 18.1 | protocol inconsistencies were recognized after three months from the start of the study - **no action** | 4 | 4 | 16 | 0 |
|  | 18.2 | Protocol Amendment revised, submitted to IRB/IEC and FDA, and distributed to sites. Monitoring Confirmation | 3 | 2 | 6 | 0 |
|  | 18.3 | Data review and correction are necessary at all sites, eight h/site | 2 | 2 | 4 | 0 |
|  | 18.4 | 100 extra monitoring hours are necessary to re-monitor changes at each site | 1 | 1 | 1 | 1 |
| **Risk** | **19** | **Inadequate monitoring revealed** | | | | |
| Mitigations | 19.1 | Inadequate monitoring revealed in one country - no action | 3 | 4 | 12 | 0 |
|  | 19.2 | A new monitor is assigned, and three re-monitoring visits/site are necessary | 1 | 1 | 1 | 1 |
| **Risk** | **20** | **Low data quality at some sites** | | | | |
| Mitigations | 20.1 | Data quality is not acceptable at five sites - no action | 4 | 4 | 16 | 0 |
|  | 20.2 | 25 extra monitoring visits are necessary | 1 | 1 | 1 | 1 |
| **Risk** | **21** | **Lack of investigator oversight at one of the sites** | | | | |
| Mitigations | 21.1 | Investigator oversight is not suitable at 20 sites - no action | 3 | 4 | 12 | 0 |
|  | 21.2 | 20 meetings with the investigators by a senior trial team member and 20 additional monitoring visits | 1 | 1 | 1 | 1 |
| **Risk** | **22** | **Risk of labeling error - manufacturing. Clinical supply is not available. Risk to study completion - time delay. It might cause new sites activation** | | | | |
| Mitigations | 22.1 | Labeling error - no action | 4 | 4 | 16 | 0 |
|  | 22.2 | Create over the label. Retain with the same manufacturer. | 3 | 2 | 6 | 0 |
|  | 22.3 | Repackage and relabel investigational drug - time delay | 2 | 1 | 2 | 1 |
| **Risk** | **23** | **Manufacturing - the batch failure rate is high** | | | | |
| Mitigations | 23.1 | Manufacturing - the batch failure rate is high - no action | 5 | 4 | 20 | 0 |
|  | 23.2 | Backup CMO#1 | 4 | 2 | 8 | 1 |
|  | 23.3 | Backup CMO #2 | 3 | 2 | 6 | 0 |
| **Risk** | **24** | **Slow drug distribution and resupply process** | | | | |
| Mitigations | 24.1 | Slow drug distribution and resupply process - no action | 3 | 4 | 12 | 0 |
|  | 24.2 | Faster delivery. Backup carrier | 3 | 2 | 6 | 1 |
|  | 24.3 | Faster delivery options + Additional depots | 2 | 2 | 4 | 0 |
| **Risk** | **25** | **Low overage. Risk of under-supply** | | | | |
| Mitigations | 25.1 | Low overage. Risk of under-supply | 4 | 4 | 16 | 1 |
|  | 25.2 | Increase overage by 5% | 2 | 3 | 6 | 0 |
|  | 25.3 | Increase overage by 10% | 2 | 2 | 4 | 0 |

Legend: 1 – mitigation option selected, 0 – not selected
